## Supplementary materials for "Modelling population genetic screening in rare neurodegenerative diseases"

**Table of contents**

1. Underlying mathematical concepts

2. Estimating analytical validity: sensitivity and specificity
3. Parameter estimates by case study
   3.1. Huntington’s disease
   3.2. amyotrophic lateral sclerosis
   3.3. phenylketonuria

**1. Underlying mathematical concepts**

The present framework follows Bayesian logic and the principles of conditional probability, which are core aspects of clinical decision making and evidence-based medicine^1^. The principles applied within this manuscript are briefly summarised below.

Bayes theorem states that

$$\begin{aligned} P\left( A | B \right)=\frac{P\left( A \right)\times P\left( B | A \right)}{P\left( B \right)}=\frac{P\left( A\cap B \right)}{P\left( B \right)} ,\#\left( S1 \right) \end{aligned}$$

letting $P$ denote probability, and $A$ and $B$ be events which have a probability of occurring independently, denoted respectively by $P(A)$ and $P(B)$, and of co-occurring at the intersection $P\left( A\cap B \right)$. The probability of event $A$ given that event $B$ has occurred is denoted as $P\left( A | B \right)$, and $P\left( B | A \right)$ represents the probability of $B$ given $A$. All events must have probabilities between 0 and 1 and the probability of not an event, denoted for not-$A$, $A’$, as $P\left( A^{'} \right)$, can be calculated by subtracting the probability of the event from the total event space (i.e. $P\left( A^{'} \right)=1-P(A)$).

We follow the principle that the total probability of a conditioned event can be derived from the probabilities of mutually exclusive occurrences of the event within the total event space. Letting $A$ represent an event conditioned upon event $B$,

$$\begin{aligned} P\left( A \right)=P\left( A | B \right)\times P\left( B \right)+P\left( A | B^{'} \right)\times P\left( B^{'} \right) .\#\left( S2 \right) \end{aligned}$$

We also apply the chain rule, which considers a third event, $C$, and states that

$$\begin{aligned} P\left( A\cap B\cap C \right)=P\left( A \right)\times P\left( B | A \right)\times P\left( C | B\cap A \right) .\#\left( S3 \right) \end{aligned}$$

Accordingly, the probability of $A$ and $B$ co-occurring given that $C$ has occurred can be derived:

$$\begin{aligned} P\left( A\cap B | C \right)=\frac{P\left( A\cap B\cap C \right)}{P\left( C \right)}=P\left( B | C \right)\times P\left( A | B\cap C \right) .\#\left( S4 \right) \end{aligned}$$

The total probability of event $A$ given the occurrence of a third event, $C$, can be determined in accordance with the principles of total probability and of equation $S4$:

$$\begin{aligned} P\left( A|C \right)=P\left( A | B\cap C \right)\times P\left( B|C \right)+P\left( A | B^{'}\cap C \right)\times P\left( B^{'} | C \right) .\#\left( S5 \right) \end{aligned}$$

If the probability of event $A$ has conditional independence from event $C$ when the occurrence or non-occurrence of event $B$ is known, then $P\left( A | B\cap C \right)=P\left( A | B\cap C^{'} \right)=P\left( A | B \right)$ and $P\left( A | B'\cap C \right)=P\left( A | B'\cap C^{'} \right)=P\left( A | B' \right)$. Thus, equation $S5$ can be simplified to:

$\begin{aligned} P\left( A|C \right)=P\left( A | B \right)\times P\left( B|C \right)+P\left( A | B^{'} \right)\times P\left( B^{'} | C \right) .\#\left( S6 \right) \end{aligned}$

**2. Estimating analytical validity: sensitivity and specificity**

Test performance parameters were defined based on the benchmarking estimates of state-of-the-art Next Generation Sequencing tools. Our framework requires estimates of the probability of a positive test result given the presence of a genetic marker, $P(T|M)$ (a.k.a. sensitivity, true positive rate, recall), and of the probability of a negative test result given the absence of a genetic marker, $P(T’|M’)$ (a.k.a. specificity, true negative rate, selectivity). Benchmarking papers we identified typically provided two performance estimates directly, $P(T|M)$ and the probability of mutation given a positive test, $P(M|T)$ (a.k.a. precision, positive predictive value). Therefore, the sensitivity was readily accessible, and we could derive specificity based on the given values of $P(T|M)$ and $P(M|T)$. To do this, we first calculated the false positive rate, $P(T|M’)$, exploiting that

$$\begin{aligned} P\left( M | T \right)=\frac{P\left( T | M \right)}{P\left( T | M \right)+P(T|M')} ,\#\left( S7 \right) \end{aligned}$$

which can be rearranged as

$$\begin{aligned} P\left( T | M^{'} \right)=\frac{P(T|M)-P\left( M | T \right)\times P(T|M)}{P\left( M | T \right)} .\#\left( S8 \right) \end{aligned}$$

This value can be used to determine specificity:

$$\begin{aligned} P\left( T’ | M’ \right)=1-P\left( T | M^{'} \right) .\#\left( S9 \right) \end{aligned}$$

Table S1 presents the best performing tools we identified for sequencing several major types of genetic variant.

| Tool | Variant type | Sensitivity | Specificity^*^ |
| --- | --- | --- | --- |
| Dragen Pipeline v3^2^ | Single nucleotide variant | 99.96% | 99.95% |
| Dragen Pipeline v3^2^ | Insertion or deletion (small) | 99.62% | 99.71% |
| ExpansionHunter^3^ | Short tandem repeat expansion | 99% | 90% |
| GRIDSS^4,5^ | Copy number variant - gene deletion | 28.9% | 95.9% |
| Wham^5^ | Copy number variant - gene duplication | 10.20% | 92.33% |
| Table S1. *Performance benchmarks of next generation sequencing tools specialised for genotyping different types of variants  ^*^Unless defined within the referenced benchmarking paper, specificity was derived via the approach detailed in the Supplementary Materials 2.* | | | |

**3. Parameter estimates by case study**

For each case study, input parameters defined within our framework were estimated using data drawn from published literature and suitable online genetic databases. Those parameters defined are:

- $P\left( D \right)$, the prior probability of the person being affected by disease $D$
- $P(M|D)$, the frequency of disease marker $M$ among those affected by disease $D$
- $P(D|M)$, penetrance, the probability of $D$ occurring for people harbouring marker $M$
- $P(T|M)$, the sensitivity (true positive rate) of the testing procedure for detecting $M$
- $P(T'|M^{'})$, the specificity (true negative rate) of the testing procedure for identifying the absence of $M$

Assumptions made across these case studies and the realities to which they correspond are shown in Table S2. Estimates of P(T|M) and P(T’|M’) were specified for each scenario of the diseases examined according to the variant type in the assessed gene which is most frequently associated with the considered disease and based on the performances reported in Table S1. Table S2 summarises the parameters assigned in each case study scenario. Below follows a description of the approach to their ascertainment.

| Assumption | Reality |
| --- | --- |
| The person undergoing genetic screening will live a normal lifespan | There is no guarantee that a person will live to the age at which a phenotype would onset |
| Analytical validity is only imperfect at the point of variant calling | Errors can be introduced at any stage of sequencing and data processing, including clerical errors, poor read quality, and incorrect alignment |
| In recessive diseases, only biallelic mutations are pathogenic and both homozygosity and compound heterozygosity result in equivalent phenotypes | Heterozygous inheritance of variants pathogenic for recessively inherited phenotypes will likely bear some consequence and compound heterozygosity may modify disease presentations |
| Variant penetrance is defined for the state of disease manifesting | A pathogenic variant may produce clinicomolecular evidence of disease in the absence of a phenotypic disease manifestation |
| Penetrance is measured only as applied to the disease named | Penetrance of variants with pleiotropic effects can be considered according to pathogenicity for any number of implicated traits |
| Table S2. *Assumptions made about the case studies described in this paper.* | |

3.1. ***Huntington’s disease (HD)***

For the blind screening scenario of the HD case study, we estimated that $P\left( D \right)=0.00041$, 1 in 2439, representing the frequency of a pathogenic *HTT* CAG short tandem repeat expansion (STRE) of ≥40 repeat units across people sampled from Scotland, the United States of America, and British Columbia^6^. We deemed this a suitable estimate of $P(D)$ because the *HTT* CAG expansion at >40 repeat units is fully penetrant within a normal lifespan and accounts for the vast majority of observed HD cases^7-9^. The estimate is also comparable to the frequency of HD cases recorded between 1986-2015 in two Norwegian death registries^10^. It is sufficiently precise for the purposes of our study.

We specified that $P(M|D)=1$, letting $M$ represent harbouring an HTT STRE of ≥40 repeat units. In reality, a small percentage of people who develop HD harbour expansions of fewer repeat units, however, as $P(D)$ is defined according to population frequency of ≥40 repeat unit *HTT* CAG expansions it would be inappropriate to define $M$ as less than 1. We similarly defined that $P(D|M)=1$, in line with the definition of $P(D)$ used in this scenario.

In the targeted testing scenario of this case study, we modelled risk for a person whose parent harbours the fully penetrant form of this $HTT$ STRE and who has a 0.5 probability of inheriting an identical variant. Therefore, we adjusted the probability of disease parameter to $P\left( D \right)=0.5$.

Sensitivity and specificity for sequencing *HTT* were based on the performance of ExpansionHunter^3^ for sequencing STREs, $P\left( T | M \right)=0.99$, $P(T’|M’)=0.90$.

3.2. ***Amyotrophic lateral sclerosis (ALS)***

In screening for ALS, $P\left( D \right)=0.0033$, 1 in 300, representing the upper-bound of estimated lifetime cumulative risk of ALS^11,12^.

We modelled several scenario of $M$ in this case study:

- For the *SOD1* (all) scenario, *M* represents harbouring any *SOD1* variant reported across the familial and sporadic ALS European population sample sets of a large meta-analysis^13^.
- For *SOD1* (A5V), harbouring the widely described *SOD1*-A5V single nucleotide variant (SNV).
- For *FUS* (all), harbouring any *FUS* variant reported across the familial and sporadic ALS European population sample sets of the previous meta-analysis^13^.
- For *FUS* (ClinVar), harbouring any of the 21 *FUS* variants recorded as pathogenic or likely pathogenic for ALS within the ClinVar Database^14^ (see Table S3).
- For *C9orf72,* harbouring a hexanucleotide, GGGGCC, STRE of ≥30 repeat units within the first intron of the *C9orf72* gene.

| *FUS* gene variant | Protein consequence | ClinVar classification | Accession |
| --- | --- | --- | --- |
| c.412_429GGACAGCAGCAAAGCTAT[1] | p.138_143GQQQSY[1] | Likely pathogenic | VCV000873229.1 |
| c.616G>A | p.Gly206Ser | Pathogenic | VCV000029708.1 |
| c.646C>T | p.Arg216Cys | Pathogenic | VCV000016227.1 |
| c.1394-2del | - | Pathogenic | VCV000447355.3 |
| c.1394-1G>T | - | Pathogenic | VCV000873230.1 |
| c.1483C>T | p.Arg495Ter | Pathogenic | VCV000029707.2 |
| c.1504_1505AG[3] | p.Gly503fs | Pathogenic | VCV000665141.1 |
| c.1509dup | p.Gly504fs | Pathogenic | VCV000933229.1 |
| c.1520G>A | p.Gly507Asp | Pathogenic | VCV000016226.1 |
| c.1540A>T | p.Arg514Trp | Likely pathogenic | VCV000803253.1 |
| c.1551C>G | p.His517Gln | Pathogenic | VCV000016221.1 |
| c.1553G>A | p.Arg518Lys | Pathogenic | VCV000016223.1 |
| c.1554_1557del | p.Gln518fs | Pathogenic | VCV001073222.1 |
| c.1555C>T | p.Gln519Ter | Pathogenic | VCV000873231.1 |
| c.1561C>T | p.Arg521Cys | Pathogenic | VCV000016224.1 |
| c.1561C>G | p.Arg521Gly | Pathogenic | VCV000016222.3 |
| c.1562G>T | p.Arg521Leu | Pathogenic | VCV000873232.2 |
| c.1562G>A | p.Arg521His | Pathogenic | VCV000016225.1 |
| c.1571G>T | p.Arg524Met | Likely pathogenic | VCV000873233.1 |
| c.1574C>T | p.Pro525Leu | Pathogenic | VCV000280110.9 |
| c.1577A>G | p.Tyr526Cys | Pathogenic | VCV000873234.1 |
| Table S3. *Variants in the FUS gene recorded in ClinVar^14^ as “pathogenic” or “likely pathogenic” for amyotrophic lateral sclerosis. ClinVar variant search performed 24/05/2021* | | | |

Estimates of $P(M|D)$ were determined for *SOD1* (all), *FUS* (all), and *C9orf72* using data from recent meta-analyses that examined the frequency of variants in these genes among people with ALS^13,15^. In these reports, variant frequencies were shown to differ between people of European and Asian ancestry and were reported separately for familial and sporadic cohorts of people with ALS, respectively representing those with and without family history of disease. We drew the variant frequency estimates reported for people of European ancestry for use in our case study. To derive the total frequency of these variants across the European ALS populations, we harmonised variant frequency estimates made in the familial and sporadic ALS sub-populations using a weighted mean calculation, where

$$\begin{aligned} P\left( M | D \right)=\left( P\left( M | D \right)_{fam}\times0.05 \right)+\left( P\left( M | D \right)_{spor}\times0.95 \right),\#(S10) \end{aligned}$$

letting $P\left( M | D \right)_{fam}$ be the variant frequency in people who have family history of ALS and $P\left( M | D \right)_{spor}$ be the variant frequency in those without family history. The weighting factors of 0.05 and 0.95 represent that approximately 5% of people with ALS have family disease history^16^. We additionally derived 95% confidence intervals for each $P(M|D)$ estimate, $P\left( M | D \right)^{95\%CI}$, by propagating the uncertainty in $P\left( M | D \right)_{fam, spor}$^17^. We first calculated the 95% margin of error, $E$, for $P\left( M | D \right)_{fam,spor}$ in the given gene. Letting $i$arbitrarily represent the familial or sporadic states and $P\left( M | D \right)_{i}^{95\%lower}$ denote the lower bound 95% interval of the $P\left( M | D \right)_{i}$ estimate,

$$\begin{aligned} E_{{P(M|D)}_{i}}=P\left( M | D \right)_{i}-P\left( M | D \right)_{i}^{95\%lower}.\#\left( S11 \right) \end{aligned}$$

These errors can then be summed in quadrature, weighted by the constants from equation S10, to obtain $E$ in $P(M|D)$:

$$\begin{aligned} E_{P(M|D)}=\sqrt{\left( E_{P{(M|D)}_{fam}}\times0.05 \right)^{2}+\left( E_{P{(M|D)}_{spor}}\times0.95 \right)^{2}},\#\left( S12 \right) \end{aligned}$$

from which confidence intervals for P(M|D) can be derived:

$$\begin{aligned} P\left( M | D \right)^{95\%CI}=P\left( M | D \right)\pm E_{P\left( M | D \right)}.\#\left( S13 \right) \end{aligned}$$

Table S4 presents our estimates of $P(M|D)$ in these three scenarios.

|  | *SOD1* (all)  [95% CI]*^13^* | *FUS* (all)  [95% CI]*^13^* | *C9orf72* (≥30 repeat units)  [95% CI]*^15^* |
| --- | --- | --- | --- |
| P(M\|D)_fam_ | 0.148 [0.115, 0.185] | 0.028 [0.021, 0.035] | 0.32 [0.28, 0.37] |
| P(M\|D)_spor_ | 0.012 [0.007, 0.019] | 0.003 [0.001, 0.005] | 0.05 [0.04, 0.06] |
| P(M\|D)^†^ | 0.0188 [0.0138, 0.0238] | 0.00425 [0.0023, 0.0061] | 0.0635 [0.0538, 0.0732] |
| Table S4. *Estimation of variant frequency among people of European ancestry with ALS, P(M\|D), for the SOD1 (all), FUS (all), and C9orf72 scenario* *of the ALS case study.*  *^†^Derived in accordance with equations S10-S13.* | | | |

For the *SOD1* (A5V) and *FUS* (ClinVar) scenarios, we estimated $P(M|D)$ using data from repositories of familial and sporadic ALS patients. The familial ALS population was represented within the ALS Variant Server^18^ and the sporadic within the Project MinE Data Browser^19^.

Seven of 1125 people with familial ALS were heterozygous for the *SOD1-*A5V variant in the ALS variant server, compared to 1 of 4366 with sporadic ALS in the Project MinE Data Browser; no people were homozygous for this variant ($P\left( M | D \right)_{fam}= 0.00622;$ $P\left( M | D \right)_{spor}=0.000229)$. Following equations S10-S13, we derived that $P\left( M | D \right)=0.000529 (95\% CI: 0, 0.0364)$ for the *SOD1* (A5V) scenario.

For the FUS (ClinVar) scenario, 8 of the 21 ALS risk variants recorded in ClinVar (Table S3) were harboured by people within the familial and sporadic databases, and just two variants occurred in both databases. Table S5 presents the frequencies of each variant in the familial and sporadic states across the two repositories; we estimated that $P\left( M | D \right)=0.00251 (95\%CI:0.000941, 0.00409)$ for the *FUS* (ClinVar) scenario.

|  | Genetic variant (protein consequence) | | | | | | | | Total  (95%CI^∆^) |
| --- | --- | --- | --- | --- | --- | --- | --- | --- | --- |
|  | **c.646C>T (p.R216C)** | **c.1483C>T (p.R495X)** | **c.1520G>A (p.G507D)** | **c.1561C>T (p.R521C)** | **c.1562G>A (p.R521H)** | **c.1562G>T (p.R521L)** | **c.1571G>T**  **(p.R524M)** | **c.1574C>T**  **(p.P525L)** |  |
| P(M\|D)_fam­_ (n/N)_­­_*^18^* | 9.950E-04  (1/1005) | 9.881E-04  (1/1012) | 0 | 7.207E-03  (8/1110) | 1.808E-03  (2/1106) | 0 | 8.905E-04  (1/1123) | 3.552E-03  (4/1126) | 1.544E-02 (1.507E-02, 1.581E-02) |
| P(M\|D)_spor_  (n/N)*^19^* | 0 | 0 | 2.290E-04  (1/4366) | 0 | 6.871E-04  (3/4366) | 4.581E-04  (2/4366) | 0 | 4.581E-04  (2/4366) | 1.832E-03  (6.261E-04, 3.038E-03) |
| P(M\|D)^†^ | 4.975E-05 | 4.941E-05 | 2.176E-04 | 3.604E-04 | 7.432E-04 | 4.352E-04 | 4.452E-05 | 6.128E-04 | **2.513E-03**  **(9.407E-04, 4.085E-03)** |
| Table S5. *Estimation of the frequency of ALS risk variants reported in the FUS gene on the ClinVar database (Table S3) among people with ALS represented in the ALS variant Server^18^ and Project MinE Data Browser^19^.*  *^†^Derived in accordance with equation S10;* ^∆^95% CI derived in accordance with equations S11-S13. | | | | | | | | | |

We estimated $P(D|M)$ for the *SOD1* (all), *FUS* (all), *FUS* (ClinVar), and *C9orf72* scenario of the ALS case study using the adpenetrance approach described in our recent publication^20^. This was selected because the modelled ALS risk variants are all rare in the population and the approach can provide population-based penetrance estimates for rare variants which avoid the ascertainment biases limiting methods which examine the distribution of a variant between affected cases and healthy controls. In the original publication describing this method, we previously estimated penetrance for some of the present case study scenarios: $P\left( D | M \right)=0.701 (95\%CI: 0.491, 0.926)$ for *SOD1* (all), and $P\left( D | M \right)= 0.439 (95\% CI: 0.358, 0.520)$ for *C9orf72*. Table S6 presents additional penetrance estimates not previously made. We estimated that $P\left( D | M \right)=0.579 (95\% CI:0.291, 0.884)$ for FUS (all), and $P\left( D | M \right)=0.538 (95\% CI: 0.282, 0.804)$ for FUS (ClinVar).

| ALS case study scenario | Variant frequency in familial state  (95%CI) | Variant frequency in sporadic state  (95% CI) | Rate of first-degree family ALS history among people with ALS | Average sibship size*^†^* | Disease states modelled^#^ | Familial disease rate among people harbouring the variant across states modelled  (95% CI) | Penetrance (95% CI) ^§^ |
| --- | --- | --- | --- | --- | --- | --- | --- |
| *-* | P(M\|D)_fam_ | P(M\|D)_spor_ | - | - | - | - | P(D\|M) |
| *FUS* (all) | 0.028 (0.021, 0.035)*^13^* | 0.003 (0.001, 0.005)*^13^* | 0.050^16^ | 1.543 | F, S | 0.329 (0.172, 0.487) | 0.579 (0.291, 0.884) |
| FUS (ClinVar) | 1.544E-02 (1.507E-02, 1.581E-02)^¥^ | 1.832E-03 (6.261E-04, 3.038E-03)^¥^ | 0.050^16^ | 1.543 | F, S | 0.307  (0.167, 0.447) | 0.538 (0.282, 0.804) |
| Table S6. *Estimation of the aggregate penetrance for ALS of FUS variants identified in people with ALS from a European population (FUS (all)), and of FUS variants identified as pathogenic for ALS within the ClinVar database (FUS (ClinVar)) following the adpenetrance approach^20^. ^†^Estimated from Total Fertility Rates reported for the European Union region in 2018*^21^; ^#^*F=familial, S=sporadic; ^¥^See Table S5;* ^§^Estimates take into account an approximated 0.0033 lifetime risk of ALS among people not harbouring the variant (denoted $\boldsymbol{g}$ within adpenetrance) – $\boldsymbol{g}$ is a conditional probability of a population member having disease given the absence of the tested variant - since the probability of the average population member having no variant in these scenarios is ~1, $\boldsymbol{g\approx P}\left( \boldsymbol{D} \right)\boldsymbol{=0.0033}$. | | | | | | | |

$P(D|M)$ was estimated as 0.91 for *SOD1* (A5V) based on figures reported previously^22^. This estimate was taken in preference to one obtained via the adpenetrance approach because of high uncertainty in the *SOD1*-A5V penetrance estimate (1 [95% CI: 0.128, 1]), reflecting its low frequency among the sporadic ALS sample; occurring in only 1 person in this sample. We note however, that the two estimates do correspond.

Sensitivity and specificity were defined for the *SOD1* (all), *SOD1* (A5V), *FUS* (all), and *FUS* (ClinVar) case studies according to the performance of the Dragen Pipeline v3^2^ for sequencing SNVs: $P(T|M)=0.9996$, and $P(T’|M’)=0.9995$. This reflects that the ALS-associated risk variants represented in these genes are predominantly SNVs^23,24^.

For the *C9orf72* marker, we modelled two testing scenarios: (1) genetic screening with sensitivity and specificity defined by performance of existing performance of ExpansionHunter^3^ for sequencing STREs, $P\left( T | M \right)=0.99$, $P(T’|M’)=0.90$. (2) using repeat-primed polymerase chain reaction with amplicon-length analysis^25^ as a secondary test to validate a positive NGS screening result from scenario 1. In the second scenario $P(D)= 0.0052$, which is the $P(D|T)$ result of scenario 1, and sensitivity and specificity are determined by performance of the secondary testing protocol^25^: $P\left( T | M \right)=0.95$, $P(T’|M’)=0.98$.

3.3. ***Phenylketonuria (PKU)***

In screening for PKU, $P\left( D \right)=0.0001$, 1 in 10,000, representing the approximate birth prevalence of PKU observed in the both the US and UK populations^26^. The disease is caused by variants in the *PAH* gene and has an autosomal-recessive inheritance pattern.

Of *PAH* genotypes associated with the occurrence of PKU, over 50% are unique to a particular person^26^ and the pathogenicity of such variants would be impossible to identify if identified within a genetic screening without further data. Accordingly, we defined $M$ as the state of being homozygous or compound heterozygous for any of the three most common *PAH* variants recorded in European populations of people with PKU, each of which is classified as pathogenic within ClinVar^14^. These *PAH* variants and their respective allele frequencies, AF, among people in Europe with PKU are: p.Arg408Trp ($AF=0.637$), c.1066-11G>A ($AF=0.11$), and p.Arg261Gln ($AF=0.11$). Their summed allele frequency is $0.857$. Per the Hardy-Weinberg equilibrium, if $q=0.857$, then $q^{2}=0.734449$, which was taken as $P(M|D)$.

We calculated $P(D|M)$ of this *PAH* marker using a Bayesian approach^27-29^, where:

$$\begin{aligned} P\left( D | M \right)=\frac{P\left( D \right)\times P\left( M | D \right)}{P\left( M \right)}=\frac{P\left( D \right)\times P\left( M | D \right)}{P\left( D \right)\times P\left( M|D \right)+\left( 1-P\left( D \right) \right)\times P(M|D^{'})},\#\left( S14 \right) \end{aligned}$$

letting $P(M)$ represent the total probability of marker $M$ and $P(M|D’)$ be the probability of $M$ among people without the disease; in rare diseases, $P(M|D')\approx P\left( M \right)$. This method was selected because the method used for the ALS case study is only suitable in autosomal dominant traits and the required input parameters can be readily derived.

$P(M|D’)$ was determined based on the allele frequencies of the three considered variants in the European (non-Finnish) population of the gnomAD v2.1.1. (control) database^30^: p.Arg408Trp ($AF= 0.002071$), c.1066-11G>A ($AF= 0.0004557$), and p.Arg261Gln ($AF= 0.0004556$). Their summed AF is $0.0029823$. Therefore, if $q= 0.0029823$, then $q^{2}=0.00000889411329$, which was taken as $P(M|D’)$. Per equation S14, this was applied alongside the previously determined estimates of $P(D)$ and $P(M|D)$ estimate $P(D|M)=0.8919914195$.

Sensitivity and specificity were defined in this case study according to the performance of the Dragen Pipeline v3^2^ for sequencing SNVs: $P(T|M)=0.9996$, and $P(T’|M’)=0.9995$.
